# Sex Disparities in Chlamydia and Gonorrhea Treatment in US Adult Emergency Departments: A Systematic Review and Meta-analysis

**DOI:** 10.1101/2024.08.20.24312317

**Authors:** Rachel E Solnick, Rahi Patel, Ethan Chang, Carmen Vargas-Torres, Maaz Munawar, Carlin Pendell, Judith E. Smith, Ethan Cowan, Keith E Kocher, Roland C Merchant

## Abstract

**Importance:** In US emergency departments (EDs), empiric antibiotic treatment for gonorrhea (GC) and chlamydia (CT) is common due to the unavailability of immediate test results. Evidence suggests sex-based disparities in treatment practices, with females potentially receiving less empiric treatment than males.

**Objective:** To investigate sex differences in empiric antibiotic treatment for GC and CT in EDs, comparing practices to subsequent laboratory-confirmed results.

**Design, Setting, and Participants:** This systematic review and meta-analysis included studies from US EDs reporting GC/CT testing and empiric antibiotic treatment from January 2010 to February 2021. A total of 1,644 articles were screened, with 17 studies (n = 31,062 patients) meeting inclusion criteria.

**Main Outcomes and Measures:** The primary outcomes were GC/CT test positivity, empiric antibiotic treatment rates, and discordance between treatment and test results, stratified by sex. Data were analyzed using a random-effects model.

**Results:** Overall GC/CT positivity was 14% (95% CI, 11%-16%): 11% (95% CI, 8%-14%) in females and 25% (95% CI, 23%-26%) in males. Empiric antibiotic treatment was administered in 46% (95% CI, 38%-55%) of cases: 31% (95% CI, 24%-37%) in females and 73% (95% CI, 65%-80%) in males. Among patients without a laboratory-confirmed infection, 38% (95% CI, 30%-47%) received treatment: 27% (95% CI, 20%-34%) of females and 64% (95% CI, 55%-73%) of males. Conversely, 39% (95% CI, 31%-48%) of patients with laboratory-confirmed infections were not treated: 52% (95% CI, 46%-57%) of females and 15% (95% CI, 12%-17%) of males.

**Conclusions and Relevance:** There is significant discordance between ED empiric antibiotic treatment and laboratory-confirmed results, with notable sex-based disparities. Females were 3.5 times more likely than males to miss treatment despite confirmed infection. These findings highlight the need for improved strategies to reduce sex-based disparities and enhance empiric treatment accuracy for GC/CT in ED settings.

**Key Points:** *Question:* Are there sex-based differences in empiric antibiotic treatment for gonorrhea and chlamydia in US emergency departments (EDs), and how do these practices compare to laboratory-confirmed results?

*Findings:* In this systematic review and meta-analysis of 17 studies with 31,062 patients, females were significantly less likely than males to receive empiric antibiotic treatment for gonorrhea and chlamydia. Additionally, 39% of patients with a laboratory-confirmed infection were not empirically treated, with females 3.5 times more likely to miss treatment than males.

*Meaning:* The findings indicate significant sex disparities in ED empiric antibiotic treatment for sexually transmitted infections, underscoring the need for improved strategies to ensure equitable and accurate treatment across sexes.

## INTRODUCTION

Sexually transmitted infections (STIs) have continued to rise in the United States (US), increasing to over 2.5 million cases of chlamydia, gonorrhea, and syphilis in 2022.^1^ Curable bacterial STIs, such as *Neisseria gonorrhea* (GC) or *Chlamydia trachomatis* (CT) left untreated can cause significant reproductive health problems for females.^2^ As government funding for STI clinics has declined,^3^ emergency departments (EDs) have seen a rise in STI-related visits and test positivity, ^4–6^ with a 39% increase from 2010 to 2013, compared to a 2% increase in non-STI ED visits.^5^

The Centers for Disease Control (CDC) recommends empiric treatment for GC and CT for patients at high risk for infection who are unlikely to return for a follow-up.^7^ However, the emergence of multi-drug resistant gonorrhea in the US^8,9^ has raised increased concerns about antibiotic stewardship.^10^

Studies have shown sex-based differences in STI treatment in EDs at a local level,^11^ with males often receiving empiric treatment more frequently than females. However, there have been no meta-analyses examining ED STI treatment rates by laboratory confirmation or sex. The last systematic review on ED STI management included data only through 2010.^12^ Given changes in STI treatment guidelines and epidemiological trends, we aimed to evaluate ED-based CT and/ or GC (hereafter, CT/GC) infection regarding test positivity, empiric treatment, discordance with lab results (potential overtreatment and undertreatment), and variation by sex.

## METHODS

We conducted a systematic review of seven databases on 2/26/2021. This systematic review and meta-analysis followed PRISMA (Preferred Reporting Items for Systematic Reviews and Meta-Analyses) guidelines and was pre-registered (PROSPERO registration: #241429). A health sciences informationist (JS) conducted searches on 2/26/2021 in the following databases: Medline (OVID), Cumulative Index to Nursing and Allied Health Literature (EBSCOHost); PsycInfo (Proquest); Cochrane Central Register of Controlled Trials (Wiley); Embase (Elsevier); Scopus (Elsevier); and Web of Science (Clarivate). Searches were performed for United States-based studies published in English from 1/1/2010 through 2/26/2021. The starting date, 1/1/2010, was selected because that date was the end date of the last known published ED STI treatment review.^12^ No other search filters were applied. The initial search was conducted in Medline and included a combination of MeSH terms (medical subject headings) and keywords in the following concept areas: 1) gonorrhea and chlamydia; 2) treatment, screening, diagnosis, testing, and antibiotics; and 3) emergency department. The other database searches were translations of the primary Medline search. Citations were exported into EndNote X9 and deduplicated. The complete searches are available in the **eMethods** of the **Supplement.**

### INCLUSION CRITERIA

Studies reporting GC or CT testing and treatment in US EDs were included. We included randomized control trials, non-randomized experimental studies, population-based observational studies, and cohorts. Only peer-reviewed publications published after 2010 were included. We did not include conference proceedings, thesis papers, abstracts, articles without original data, studies with incomplete data on STI treatment, and studies for which the infection was not laboratory-confirmed. We also excluded papers with only pediatric ED populations.

### ARTICLE SCREENING

To conduct article screening, we uploaded publications from the electronic search to DistillerSR software. The citations were screened independently by a minimum of two screeners from the author team (EC, MM, CP, EC), according to the following levels of screening: 1) Abstract involved GC/CT and US Adult EDs; 2) Study manuscript included data on GC/CT prevalence and treatment, and GC/CT was tested using a nucleic acid amplification testing (NAAT). Disagreements were resolved in consensus with the study’s senior author (RS).

### DATA EXTRACTION

We used Microsoft Excel to prepare data extraction sheets. Three reviewers (MM, EC, RP) independently extracted the data and completed the sheets. Then, the data extraction results were compared, and disagreements were resolved in consensus with the study’s senior author.

Our main outcomes were proportions defined as follows: **A)** Confirmed GC/CT among Tested, i.e., test positivity: (laboratory-confirmed positive patients)/ (all tested patients); **B)** Empiric Antibiotic Treatment: treated patients/ all tested patients; **C)** Antibiotic Treatment in Patients without Confirmed GC/CT, i.e. potential overtreatment: (treated patients with laboratory-confirmed negative)/ (all patients laboratory-confirmed negative); **D)** Antibiotic Treatment Not Provided among patients with confirmed GC/CT i.e. potential undertreatment: (untreated patients with laboratory-confirmed positive) / (all patients laboratory-confirmed positive).

If data from the published manuscript required for data extraction were unavailable, the study’s first authors were emailed to request the missing data. We manually searched the references of a previously published STI treatment in the ED review article^12^ and the references of the final 17 articles to ensure our search strategy included all relevant articles. This process did not identify any additional articles that fit our inclusion criteria.

### RISK OF BIAS

Two reviewers independently assessed bias using a risk of bias tool modified from the Joanna Briggs Institute (JBI) Critical Appraisal Tool.^13^ Disagreements were resolved in consensus with a third study team member. A threshold of 5 out of 7 was used to determine if the study was of sufficient quality to be included in the study.

### STATISTICAL ANALYSIS

Estimated pooled laboratory-confirmed GC/CT test positivity prevalences and antibiotic treatment proportions were calculated from the extracted data. We calculated the 95% percent confidence intervals (CIs) for these estimates using exact binomial confidence intervals. Freeman-Tukey double arcsine transformation was used to stabilize the variance of pooled proportion estimates.^14^ Subgroup meta-analyses were conducted based on sex. Where available, estimates are reported pooled by sex; when sex was not specified in the extracted data from articles, estimates are presented as a “combined sex” group. We used a random-effects mode due to anticipated data heterogeneity. The heterogeneity of the included studies was assessed with the I² tests. An I² value of greater than 50% is indicative of significant heterogeneity. All p-values were two-tailed, and statistical significance was P<0.05. All statistical analyses were performed using Stata 16 (College Station, TX: StataCorp LLC) with the Metaprop_onecommand.^15^

## RESULTS

All 17 studies screened using the risk of bias JBI Tool were included (**eTable 1** for Risk of Bias tool). The database search identified 1644 articles included for screening. Of these, 197 were included for full-text review. Seventeen studies comprising 31,062 ED patients tested for STIs met inclusion criteria and were included in the final analysis. (**see Figure 1)**

**Figure 1.**
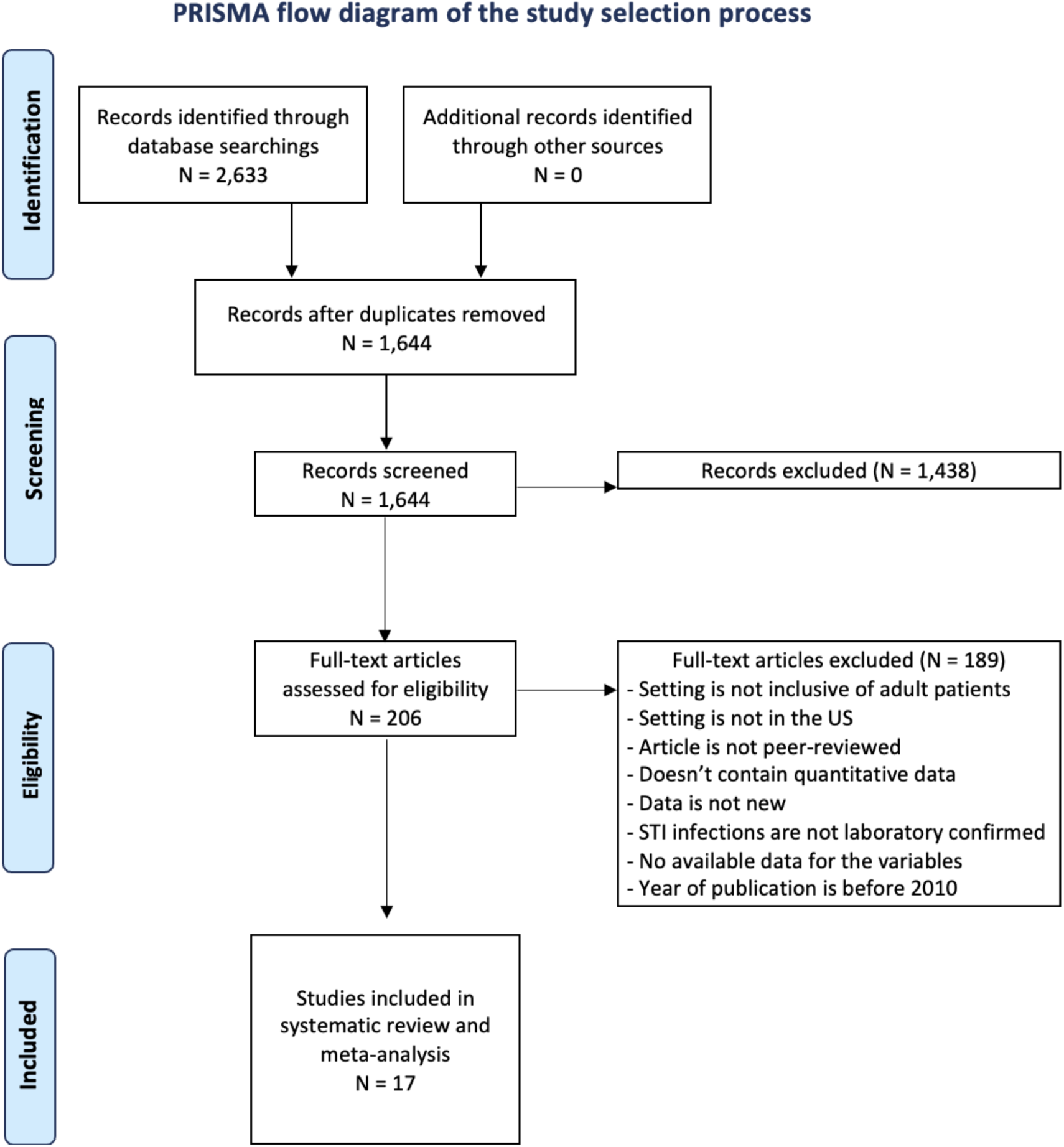
PRISMA Diagram for Study Inclusion.

**Table 1** summarizes patient demographic characteristics, treatment frequencies, and criteria met by each article. **Table 1** also provides data on included studies, all deemed to be of sufficient methodological quality as determined by the Joanna Briggs Institute tool (i.e., a score of ≥ 5)

**Table 1.**
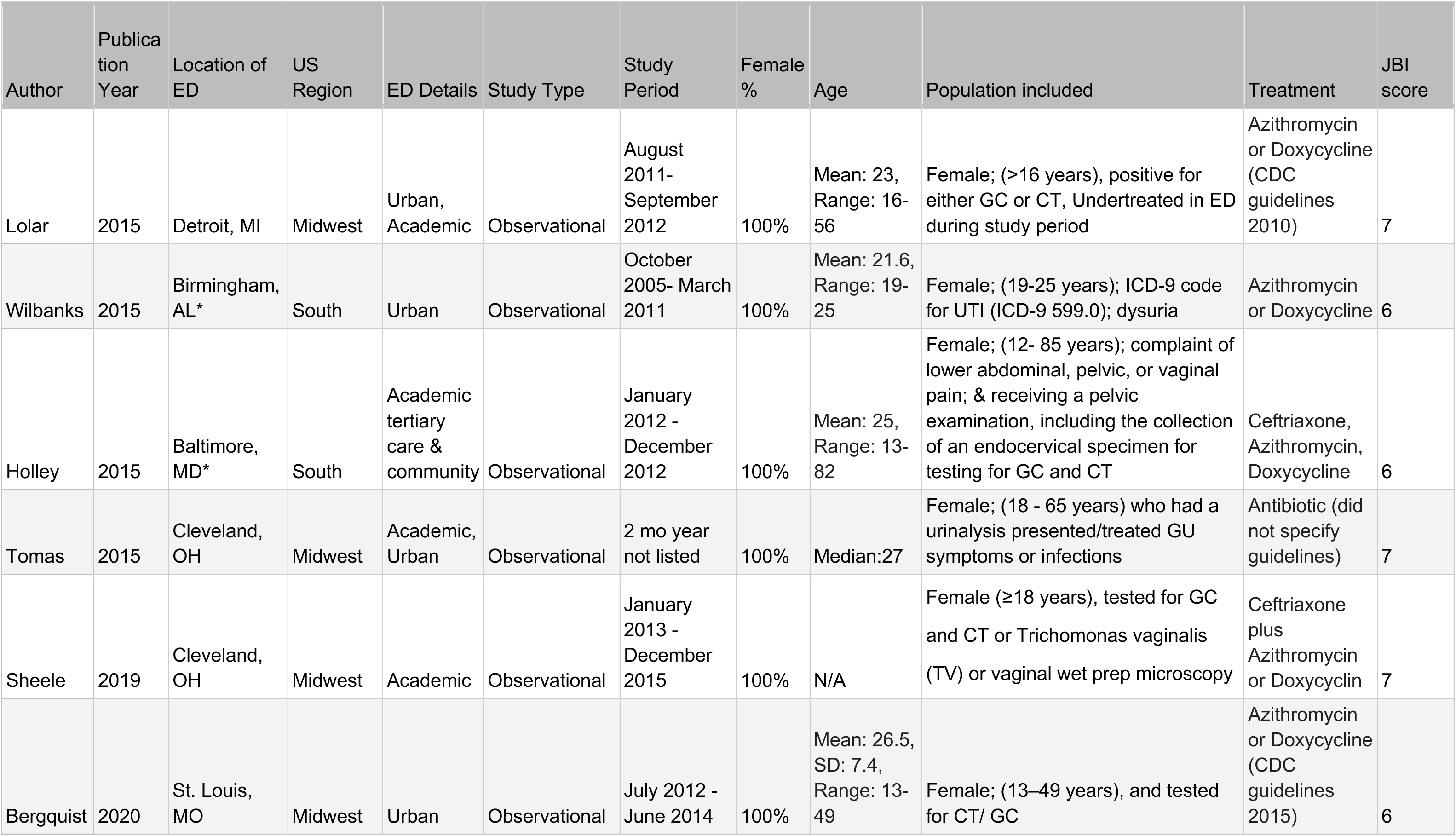
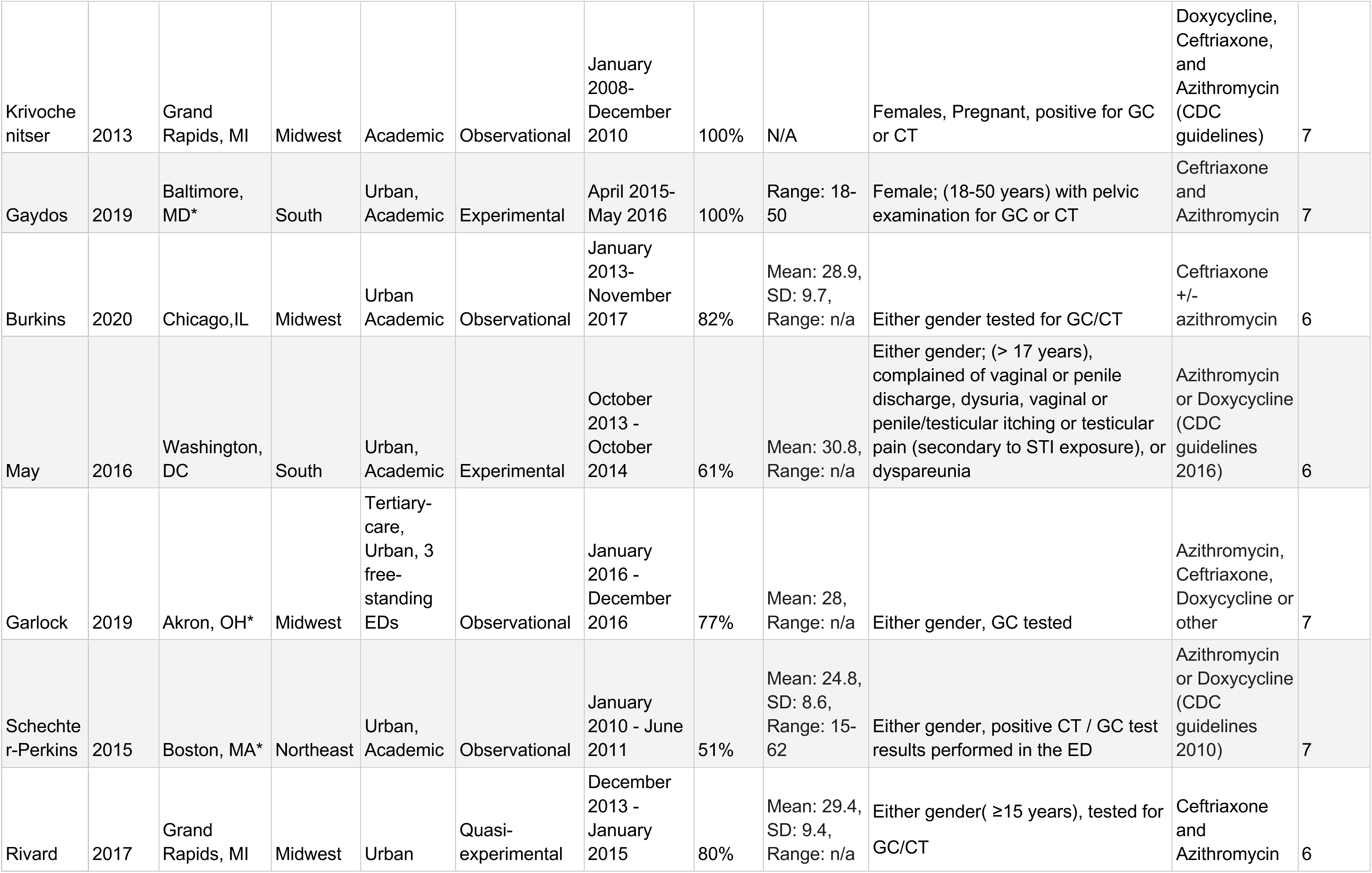
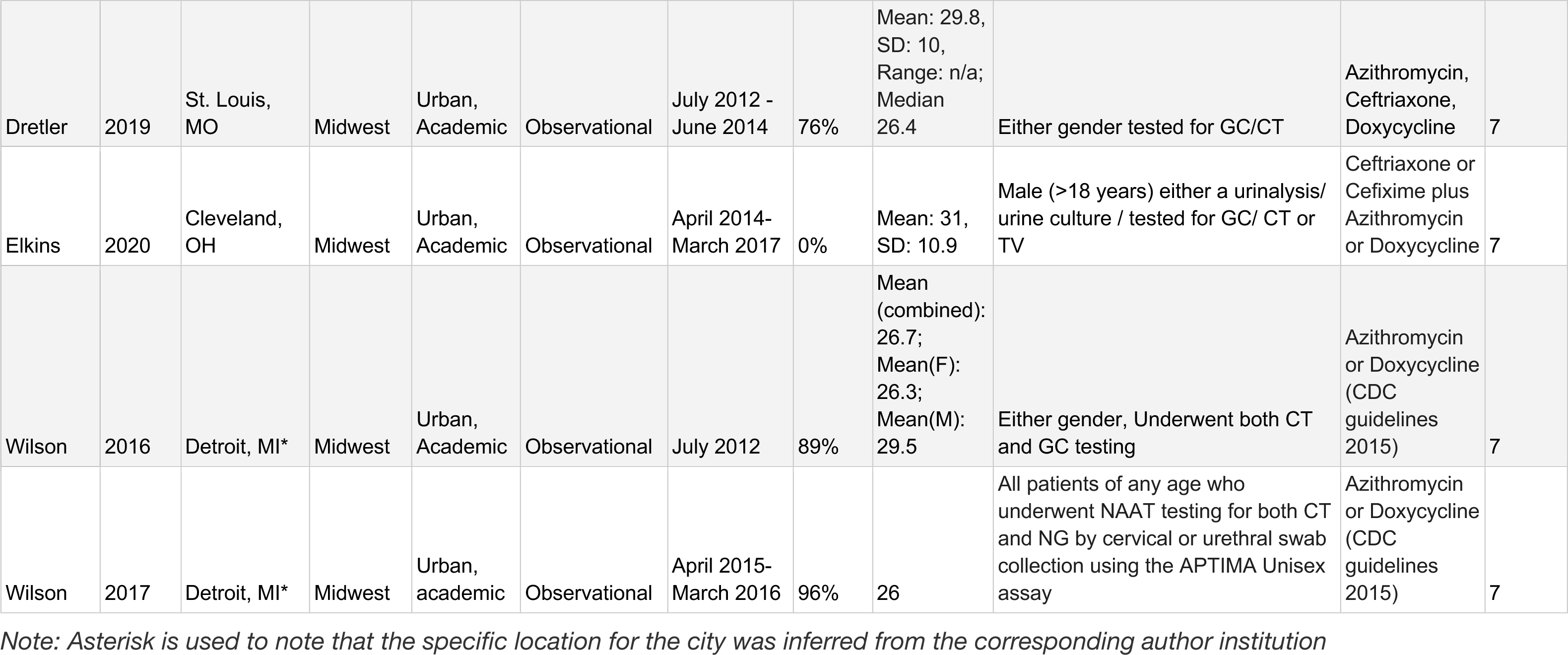
Descriptive Characteristics of Included Studies and the Risk-of-Bias Assessment Using the Joanna Briggs Institute Tool.

Among the included studies, eight (47%) included only female patients, eight (47%) both female and male patients, and one (6%) male patients only. Four (29%) were in urban locations, three (18%) were specified as occurring in academic hospitals, and 10 (59%) were in urban and academic locations. Six (35%) studies were published in 2015 or before. Eleven (65%) studies were published in 2016 or later. Six (35%) studies had a population size of less than 500, and 11 (65%) studies had a population size greater than 500.

### STI Test Results and Treatment

The forest plots for the pooled prevalence of STI testing and treatment by sex subgroups are shown in Figures 2 through 5. For a tabular form specifying the total cases of individuals, see **eTable 2.**

**Figure 2.**
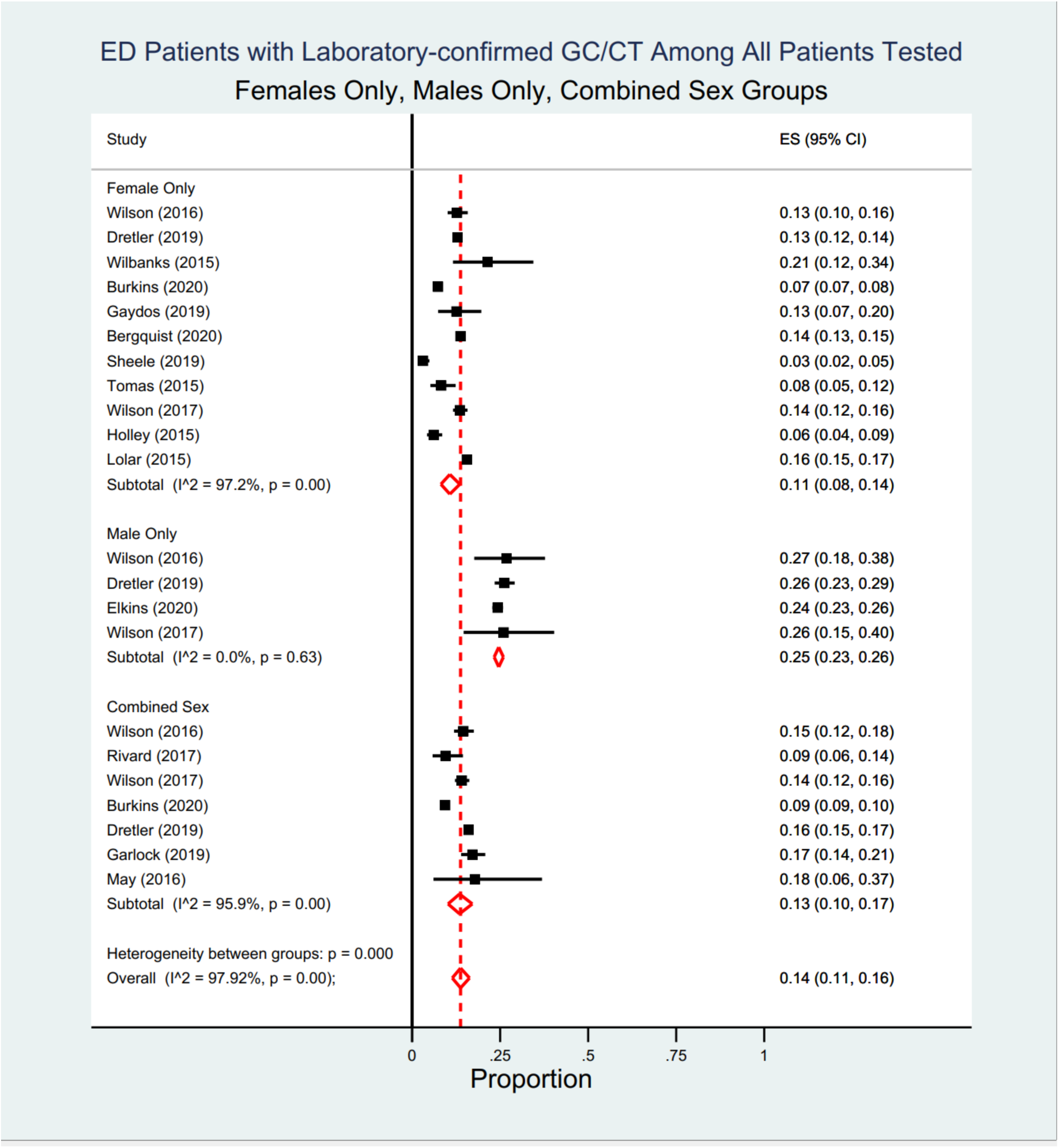
GC/CT Infection Positivity: Prevalence of Laboratory-confirmed GC/CT in ED patients Tested. *Note: Proportion refers to the following: (laboratory-confirmed positive patients)/ (all tested patients)*

#### A. Confirmed GC/CT Among Tested, i.e. Test Positivity

Figure 2 shows the pooled prevalence for laboratory-confirmed GC/CT among all ED patients tested. The overall pooled prevalence of GC/CT was 14% (95% CI, 11%-16%). Subgroup analysis shows the prevalence was significantly lower among females, at 11% (95% CI, 8%-14%), compared with males, who had a prevalence of 25% (95% CI, 23%-26%). Thus, females were approximately two-fold less likely than males to have laboratory-confirmed GC/CT.

#### B. Empiric Antibiotic Treatment

Figure 3 shows the forest plot for the pooled proportions by sex for ED patients empirically treated for GC/CT among those tested. The overall pooled proportion of patients receiving empiric antibiotic treatment for GC/CT was 46% (95% CI, 38%-55%). Subgroup analysis by sex revealed that the proportion was significantly lower in females, at 31% (95% CI, 24%-37%), compared with males, who had a proportion of 73% (95% CI, 65%-80%). This suggests that females were approximately two-fold less likely than males to receive empiric antibiotic treatment for GC/CT.

**Figure 3.**
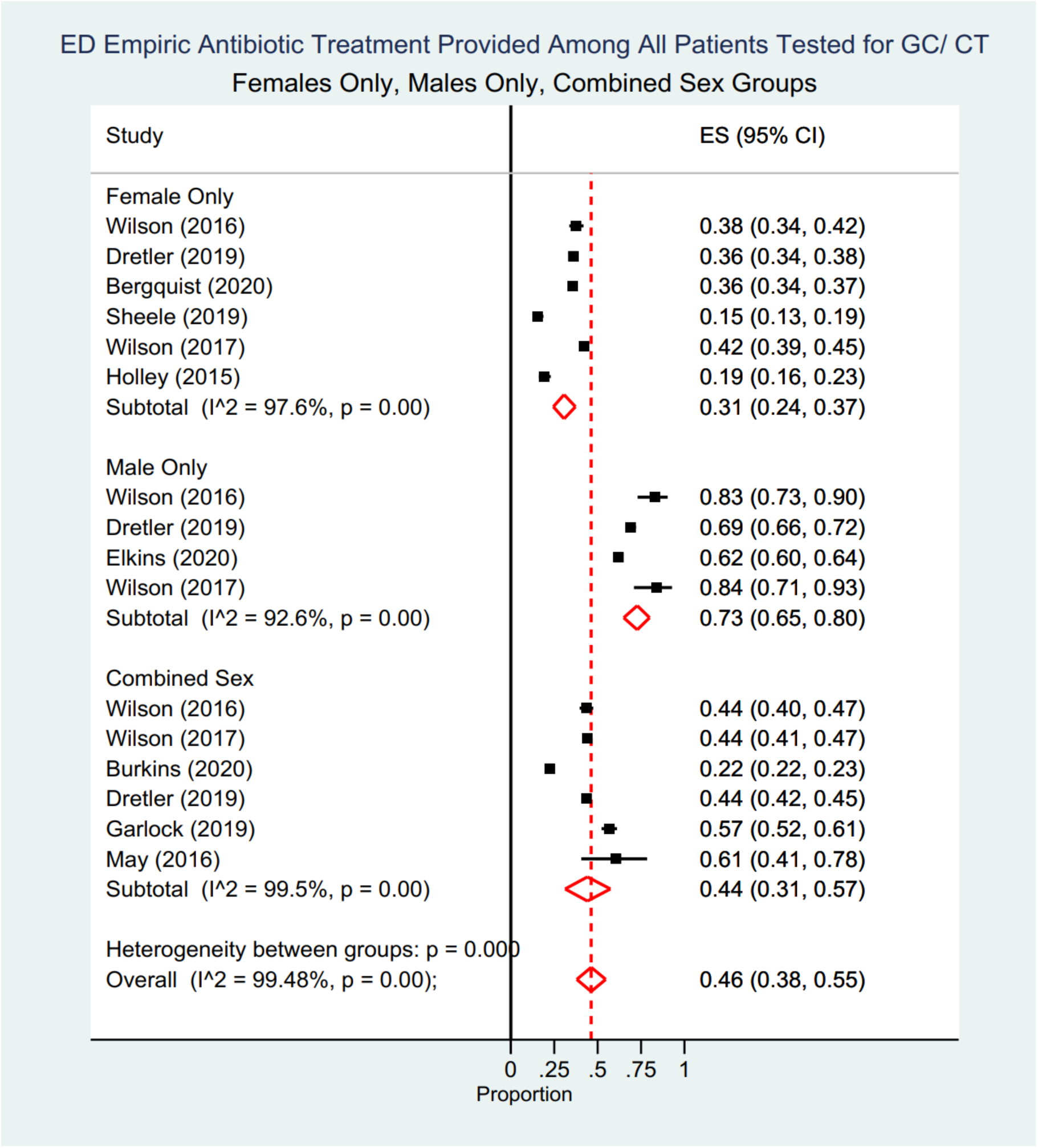
Empiric Antibiotic Treatment in all ED Patients Tested for GC/CT. *Note: Proportion refers to the following: treated patients/ all tested patients*

#### C. Antibiotic Treatment in Patients without Confirmed GC/CT, i.e. Potential Overtreatment

Figure 4 presents the forest plot of pooled proportions by sex for ED patients empirically treated for GC/CT among those without a laboratory-confirmed diagnosis. The overall pooled proportion of patients receiving empiric antibiotic treatment for GC/CT despite lacking a laboratory-confirmed diagnosis was 38% (95% CI, 30%-47%). Subgroup analysis showed that the proportion was significantly lower in females, at 27% (95% CI, 20%-34%), compared with males, who had 64% (95% CI, 55%-73%). This indicates that females were approximately two-fold less likely than males to receive empiric antibiotic treatment for GC/CT without a laboratory-confirmed diagnosis.

**Figure 4.**
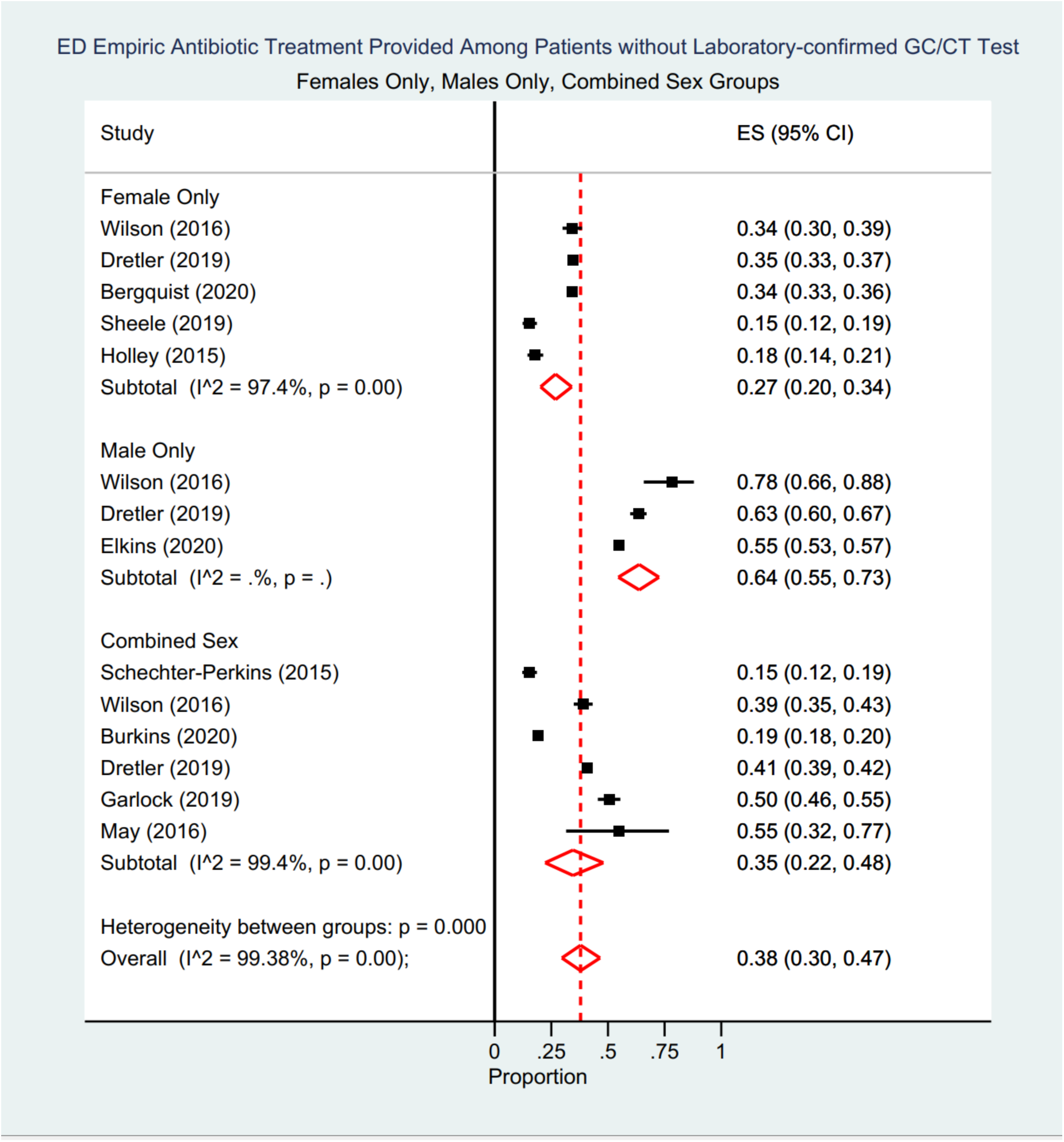
Empiric Antibiotic Treatment in ED Patients Without Laboratory-confirmed GC/CT. *Note: Proportion refers to the following: (treated patients with laboratory-confirmed negative results)/ (all patients laboratory-confirmed negative)*

#### D. Antibiotic Treatment Not Provided Among Patients with Confirmed GC/CT, i.e. Potential Undertreatment

Figure 5 shows the forest plot of pooled proportions by sex for ED patients with laboratory-confirmed GC/CT diagnoses who did not receive empiric antibiotic treatment. The overall pooled proportion of patients with a laboratory-confirmed diagnosis who were not empirically treated was 39% (95% CI, 31%-46%). Subgroup analysis revealed that this proportion was significantly higher in females, at 52% (95% CI, 46%-57%), compared with males, who had a proportion of 15% (95% CI, 12%-17%). This indicates that females were approximately 3.5-fold more likely than males to not receive empiric antibiotic treatment despite having a laboratory-confirmed GC/CT diagnosis.

**Figure 5.**
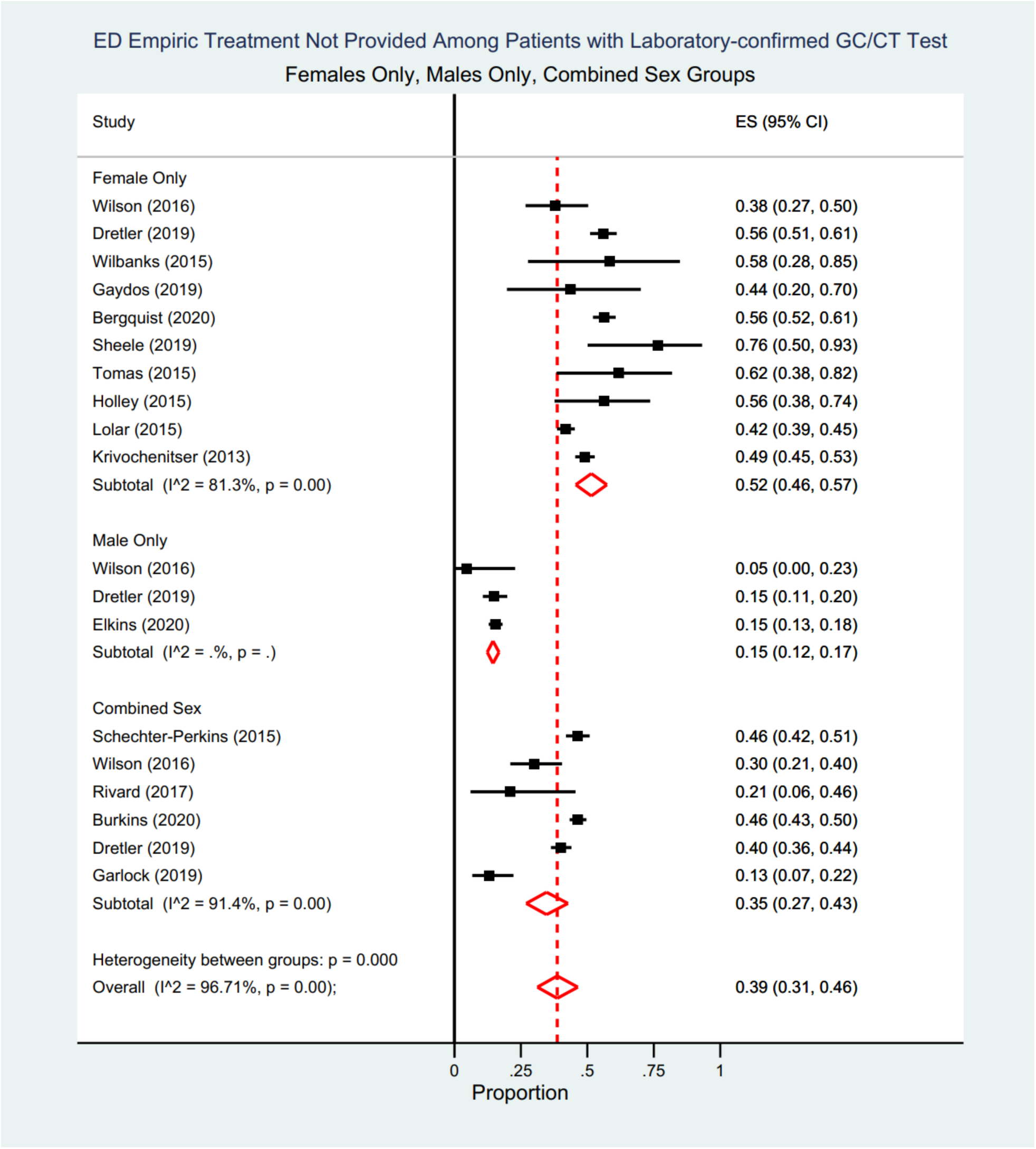
Empiric Antibiotic Treatment Not Provided among ED Patients with Laboratory-confirmed GC/CT. *Note: Proportion refers to the following: (untreated patients with laboratory-confirmed positive results) / (all patients laboratory-confirmed positive)*

## DISCUSSION

This GC/CT treatment meta-analysis in US EDs reveals substantial discordance between empiric treatment practices and final laboratory results. Results also demonstrate significant sex differences in empiric treatment, with females more frequently undertreated. Females tested for GC/CT were half as likely to receive empiric antibiotics and 3.5 times more likely to be undertreated than males. These findings underscore critical sex disparities in empiric antibiotic treatment in EDs, particularly highlighting the need for targeted interventions to address the undertreatment of females for STIs.

This meta-analysis builds on previous findings of discordance between empiric treatment and laboratory results in EDs. A 2013 database review of 1,877 visits by females aged 14-21 at an academic ED found that over half were potentially overtreated based on clinical judgment.^16^ Our findings show a slightly lower rate of potential overtreatment (27%), likely due to the inclusion of older age groups with lower STI incidence.^17^ The most recent comprehensive review of ED STI treatment before this meta-analysis, published in 2013, found wide variation in potential undertreatment at 38%–84% of general patients.^12^ We found overall potential undertreatment at the lower end of the spectrum at 39%, which may reflect a change in practice, such that there has been an increase^5^ in patients receiving treatment for STIs versus a more narrow screening selection.

Sex differences in empiric antibiotic treatment for GC/CT may stem from epidemiologic trends, disease etiology, and provider biases. Men, particularly gay and bisexual men,^18^ have the highest rates of GC and syphilis.^19^ They are more likely to present with symptomatic GC^20^, potentially leading to higher empiric treatment rates, as demonstrated in our results. Conversely, women may be more frequently tested for GC/CT in the ED due to a broader differential diagnosis in patients with lower abdominal or pelvic complaints but have lower test positivity rates and are often asymptomatic,^21,22^ which may reduce empiric treatment. Implicit and explicit gender biases^23^ may also drive these disparities.^24^ For instance, an ED prospective cohort study found that women were up to 25% less likely to receive opioids for acute abdominal pain and waited longer to receive analgesia.^25^ Addressing biases through provider self-assessment could facilitate improved patient interactions and treatment equity.^26^

Despite the rationale for these sex-based treatment differences, sex disparities in sexual and reproductive healthcare have negative consequences for women. Untreated females face an increased risk of HIV and adverse pregnancy outcomes, including spontaneous abortion, preterm birth, and infant death.^27^ Thus, the grave health reproductions for females and potential future pregnancies deserve attention and mitigation from the negative impact of empiric antibiotic treatment differences.

### PRACTICE AND POLICY IMPLICATIONS AND SOLUTIONS

Developing a shared decision-making (SDM) approach could mitigate the potential overtreatment of STIs in EDs. SDM is already employed in the ED for situations such as CT scans for renal colic.^28^ In patients with reliable contact mechanisms after the ED visit, SDM could be used with clinical gestalt to decide whether to empirically treat or defer until laboratory confirmation via routinely provided telephone “callback.” Studies reporting ED telephone callback success ranged between 53%-93%, ^29,30,31,32,33^ and reduced STI undertreatment from an average of 67% to 16%,^12^ suggesting the viability of the follow-up approach. Future research could investigate SDM’s role in ED antibiotic stewardship for STIs.

Implementing rapid diagnostic tests (RDTs) in EDs could reduce discordance between empiric treatment and laboratory results. RDTs yield test results during the ED visit and improve the clinical accuracy of treatment, decrease antimicrobial usage, and reduce ED return rates.^34–36^ While RDTs can cost an ED more than a standard laboratory test, they can potentially reduce overall costs through less staff time for follow-up. While most EDs lack RDTs for GC/CT, new technologies offer opportunities for expansion.^37^ Future studies could investigate the integration of RDTs in EDs over time with its relationship to empiric antibiotic treatment.

The high proportion of ED patients potentially undertreated presents an opportunity to offer more comprehensive STI care during a “callback.” During this telephone call, an ED clinician typically reports the positive STI finding and encourages the patient to return to the ED for treatment. Another opportunity during this call is STI partner management. Expedited partner therapy (EPT), the evidence-based^38^ practice of treating unseen partners of patients with STIs, is little used in EDs.^39^ Given that EPT has only recently become allowed or permissible in every US state,^40^ offering EPT in an ED callback for STIs is a readily attainable intervention to increase treatment and reduce STI reinfections. Additionally, HIV prevention medication, PrEP, has recently been recommended by the Centers for Disease Control for an expanded number of populations and indications, with STIs being a recognized risk factor for HIV seroconversion.^41^ This STI callback setting also presents a natural opportunity for EDs to consider PrEP education and initiation in settings with good linkage to community partners, especially considering the underrecognized role EDs can play in HIV prevention.^42^

### STRENGTH AND LIMITATIONS

The strength of this meta-analysis lies in the extensive data set of high-quality clinical data, encompassing a large cohort of individuals tested and treated across various EDs in the US. Additionally, the study addresses sex-specific differences in empiric treatment, a critical gap in the existing literature on STI management in ED settings. However, this study has limitations. First, this analysis is limited to treatment administered during the ED visit, without data on follow-up treatments (e.g., via telephone callbacks), which may reduce the apparent undertreatment rates. In patients without laboratory-confirmed infection, recent exposure may have occurred within the window before detection, making empiric treatment appropriate even if not confirmed by lab results. Second, many of the articles included did not provide sex-specific results on GC/CT prevalence or treatment, limiting our ability to analyze sex differences across all studies. Third, data did not differentiate between genital and extragenital GC/CT diagnoses, which is crucial for managing STIs, especially among MSM.^43,44^ Lastly, some studies^11,29,30^ included patients with unknown symptom status, which could influence the decision to provide empiric treatment, particularly in asymptomatic cases..^45,46^

### CONCLUSION

In this meta-analysis of adult patients tested for GC/CT in US EDs, we found significant sex differences in empiric antibiotic treatment and large discordances between empiric antibiotic treatment and laboratory test results. These findings highlight an urgent need to develop strategies to address sex disparities and improve the accuracy of empiric antibiotic treatment for GC/CT in ED settings.

## Financial disclosure area/competing interest

none

## Disclaimer

The views expressed in this article are those of the authors and do not reflect the official position of the National Institutes of Health of the University of Michigan.

## Data Availability

All data produced in the present work are contained in the manuscript

## SUPPLEMENT

**eTable 1.** RISK OF BIAS JBI TOOL.

|  | Yes | No | Unclear | Not applicable |
| --- | --- | --- | --- | --- |
| 1. Was the sample frame appropriate to address the target population? (Was it adult ED patients being tested for STIs?) |  |  |  |  |
| 2. Were the criteria for inclusion in the sample clearly defined? (i.e., how were patient selected) | ☐ | ☐ | ☐ | ☐ |
| 3. Were the study subjects and the setting described in detail (i.e. did they describe details of the ED?) | ☐ | ☐ | ☐ | ☐ |
| 4. Was the STI measured in a valid and reliable way (i.e. NAAT)? | ☐ | ☐ | ☐ | ☐ |
| 5. Were the outcomes (antibiotic treatment) measured in a valid and reliable way? (i.e., did they specify what constituted antibiotic treatment?) | ☐ | ☐ | ☐ | ☐ |
| 6. Was appropriate statistical analysis used? (i.e. Did they list enough data to get the numerator and denominator for prevalence or antibiotic treatment in lab-result positive or lab-result negative patients?) | ☐ | ☐ | ☐ | ☐ |
| 7. Was appropriate statistical analysis used? (i.e. Did they list enough data to get the numerator and denominator for prevalence or antibiotic treatment in lab-result positive or lab-result negative patients?) | ☐ | ☐ | ☐ | ☐ |
| 8. Was the sample size adequate? (i.e. was it at least 92 people?) |  |  |  |  |

Overall appraisal: Include ☐ Exclude ☐ Seek further info ☐
Comments (Including reason for exclusion)

**eTable 2.** Tabular form of data regarding treatment and lab-confirmation status.

Overall appraisal: Include ☐ Exclude ☐ Seek further info ☐ Comments (Including reason for exclusion)
| Author | Year | Female percentage | Tested | Treated with antibiotics | Negative and treated (Overtreat Numerator) | Total negative (Overtreat denominator) | Positive and not treated (Undertreat numerator) | Total positive (Undertreat denominator ) |
| --- | --- | --- | --- | --- | --- | --- | --- | --- |
| Krivochenitser | 2013 | 100% |  |  |  |  | 361 | 735 |
| Holley | 2015 | 100% | 522 | 101 | 87 | 490 | 18 | 32 |
| Lolar | 2015 | 100% | 5536 |  |  | 4675 | 361 | 861 |
| Tomas | 2015 | 100% | 254 |  |  | 233 | 13 | 21 |
| Wilbanks | 2015 | 100% | 56 |  |  | 44 | 7 | 12 |
| Schechter-Perkins | 2015 | 51% |  |  | 76 | 500 | 232 | 500 |
| Wilson | 2016 | 0% | 82 | 68 | 47 | 60 | 1 | 22 |
| Wilson | 2016 | 100% | 557 | 210 | 166 | 486 | 27 | 71 |
| wilson | 2016 | 87% | 639 | 278 | 213 | 546 | 28 | 93 |
| May | 2016 | 61% | 28 | 17 | 11 | 20 |  | 5 |
| Rivard? | 2017 | 76% | 200 |  |  | 181 | 4 | 19 |
| Dretler | 2019 | 0% | 950 | 657 | 445 | 701 | 37 | 249 |
| Dretler | 2019 | 100% | 3057 | 1097 | 924 | 2663 | 221 | 394 |
| Dretler | 2019 | 76% | 4007 | 1754 | 1369 | 3364 | 258 | 643 |
| Gaydos | 2019 | 100% | 127 |  |  | 111 | 7 | 16 |
| Sheele | 2019 | 100% | 549 | 85 | 81 | 530 | 13 | 17 |
| Garlock | 2019 | 89% | 490 | 278 | 205 | 406 | 11 | 84 |
| Elkins | 2020 | 0% | 3132 | 1945 | 1301 | 2371 | 117 | 761 |
| Bergquist | 2020 | 100% | 3908 | 1388 | 1154 | 3371 | 303 | 537 |
| Burkins | 2020 | 100% | 8591 |  |  | 7959 |  | 632 |
| Burkins | 2020 | 82% | 10452 | 2334 | 1809 | 9471 | 456 | 981 |
| Wilson | 2017 | 100% | 1112 | 470 |  |  |  | 151 |
| Wilson | 2017 | 96% | 1162 | 512 |  |  |  | 164 |
| Wilson | 2017 | 0% | 50 | 42 |  |  |  | 13 |

*Notes on extracted data:*
- Dretler et al., Wilson 2016 and 2017 et al.: We extracted data by sex group & combined sex data. - May et al., Gaydos et al., Rivard et al.: Papers included data from rapid testing approaches with immediate test results (vs. standard NAAT test with results after ED visit). We extracted only control group values to maintain consistency with the rest of the papers that used standard NAAT testing. - Gaydos et al.: Paper CT, GC, and co-infected patients separately. We prevented double counting co-infected patients by adding the individual CT and GC cases and then subtracting cases of co-infection. Co-infected cases were treated only if they received treatment for both CT and GC. - Tomas et al., the authors considered treatment after ED discharge in their total numbers for treatment. To maintain consistency with other papers that only included empiric treatment while in the ED, we did not include Tomas et al.'s treatment data.

### eMethods Search Criteria

#### Medline

**Date searched:** February 26, 2021

**Results:** 415

**Dates:** 2010-date of search

#1:

exp Chlamydiaceae Infections/ or Chlamydia trachomatis/ or Chlamydia/ or Chlamydiaceae/ or Gonorrhea/ or Neisseria gonorrhoeae/ or Sexually Transmitted Diseases/ or Sexually Transmitted Diseases, Bacterial/ or (gonococ* or gonorrh* or trachomatis or chlam?di*).tw,kf. or (sexually transmitted adj2 (disease* or infection*)).tw,kf. or (STD or STDs or STI or STIs).tw,kw.

#2:

Medical Overuse/ or drug therapy.fs. or therapy.fs. or exp diagnosis/ or mass screening/ or (treat* or therap* or overtreat* or undertreat* or over-treat* or under-treat* or screen* or diagnos* or detect* or test or tested or testing).tw,kw. or exp Anti-Bacterial Agents/ or ceftriaxone/ or azithromycin/ or doxycycline/ or (antibiot* or anti-bacter* or antibacter* or antimicrobial or anti-microbial or ceftriaxone or azithromycin or doxycycline).tw,kf.

#3:

Exp Emergency Service, hospital/ or emergenc*.ti. or (emergenc* adj2 (depart* or room* or service* or unit* or ward or wards)).tw,kw.

#4

(#1 AND #2 AND #3)

#### Scopus

**Date searched:** February 26, 2021

**Results:** 524

Dates: 2010-date of search

**#1:**

TITLE-ABS-KEY (gonococ* OR gonorrh* OR trachomatis OR chlam?di* OR (“sexually transmitted” W/2 (disease* OR infection*)) OR std OR stds OR sti OR stis)

#2:

INDEXTERMS(“Medical Overuse”) OR TITLE-ABS-KEY((treat* OR therap* OR overtreat* OR undertreat* OR over-treat* OR under-treat* OR screen* OR diagnos* OR detect* OR test OR tested OR testing OR antibiot* OR anti-bacter* OR antibacter* OR antimicrobial OR anti-microbial OR ceftriaxone OR azithromycin OR doxycycline))

#3:

INDEXTERMS(“Emergency Service, hospital”) OR TITLE-ABS-KEY((emergenc* W/2 (depart* OR room* OR service* OR unit* OR ward or wards))

)

#4

(#1 AND #2 AND #3)

#### CINAHL

**Date searched**: February 26, 2021

**Results:** 320

**Dates:** 2010-date of search

#1:

(MH “Chlamydiaceae Infections+”) OR (MH “Chlamydiaceae+”) OR (MH “Gonorrhea”) OR (MH “Neisseria”) OR (MH “Sexually Transmitted Diseases”) OR (MH “Sexually Transmitted Diseases, Bacterial”) OR ((TI gonococ* OR AB gonococ*) OR (TI gonorrh* OR AB gonorrh*) OR (TI trachomatis OR AB trachomatis) OR (TI chlam#di* OR AB chlam#di*)) OR ((TI “sexually transmitted” OR AB “sexually transmitted”) N2 ((TI disease* OR AB disease*) OR (TI infection* OR AB infection*))) OR ((TI STD OR AB STD) OR (TI STDs OR AB STDs) OR (TI STI OR AB STI) OR (TI STIs OR AB STIs))

#2:

MW “drug therapy” OR MW therapy OR (MH “diagnosis”) OR ((TI treat* OR AB treat*) OR (TI therap* OR AB therap*) OR (TI overtreat* OR AB overtreat*) OR (TI undertreat* OR AB undertreat*) OR (TI over-treat* OR AB over-treat*) OR (TI under-treat* OR AB under-treat*) OR (TI overuse OR AB overuse) OR (TI screen* OR AB screen*) OR (TI diagnos* OR AB diagnos*) OR (TI detect* OR AB detect*) OR (TI test OR AB test) OR (TI tested OR AB tested) OR (TI testing OR AB testing)) OR (MH “Antibiotics”) OR (MH “ceftriaxone”) OR (MH “azithromycin”) OR (MH “doxycycline”) OR ((TI antibiot* OR AB antibiot*) OR (TI anti-bacter* OR AB anti-bacter*) OR (TI antibacter* OR AB antibacter*) OR (TI antimicrobial OR AB antimicrobial) OR (TI anti-microbial OR AB anti-microbial) OR (TI ceftriaxone OR AB ceftriaxone) OR (TI azithromycin OR AB azithromycin) OR (TI doxycycline OR AB doxycycline))

#3:

(MH “Emergency Service”) OR TI emergenc* OR ((TI emergenc* OR AB emergenc*) N2 ((TI depart* OR AB depart*) OR (TI room* OR AB room*) OR (TI service* OR AB service*) OR (TI unit* OR AB unit*) OR (TI ward OR AB ward) OR (TI wards OR AB wards)))

#4

(#1 AND #2 AND #3)

#### Embase

**Date searched**: February 26, 2021

**Results:** 596

**Dates:** 2010-date of search

#1:

’Chlamydiaceae Infection’/exp OR ‘Chlamydiaceae’/exp OR ‘Gonorrhea’/de OR ‘Neisseria gonorrhoeae’/de OR ‘Sexually Transmitted Disease’/de OR (gonococ* OR gonorrh* OR trachomatis OR chlam?di*):ti,ab,kw OR (“sexually transmitted” NEAR/2 (disease* OR infection*)):ti,ab,kw OR (STD OR STDs OR STI OR STIs):ti,ab,kw

#2:

’overdiagnosis’/de OR ‘drug therapy’/de OR ‘therapy’/de OR ‘diagnosis’/exp OR ‘screening’/de OR (treat* OR therap* OR overtreat* OR undertreat* OR over-treat* OR under-treat* OR screen* OR diagnos* OR detect* OR test OR tested OR testing):ti,ab,kw OR ‘Antibiotic Agent’/de OR ‘ceftriaxone’/de OR ‘azithromycin’/de OR ‘doxycycline’/de OR (antibiot* OR anti-bacter* OR antibacter* OR antimicrobial OR anti-microbial OR ceftriaxone OR azithromycin OR doxycycline):ti,ab,kw

#3

’Emergency ward’/de OR emergenc*:ti OR (emergenc* NEAR/2 (depart* OR room* OR service* OR unit* OR ward OR wards)):ti,ab,kw

#4

AND [embase]/lim NOT ([embase]/lim AND [medline]/lim)

#5

(#1 AND #2 AND #3 AND #4)

#### Web of Science*

**Date searched:** February 26, 2021

**Results:** 573

*There was no date filter for this search. The date range was inadvertently left off.

#1:

TS=(gonococ* OR gonorrh* OR trachomatis OR chlam?di* OR (“sexually transmitted” NEAR/2 (disease* OR infection*)) OR STD OR STDs OR STI OR STIs)

#2:

TS=(“Medical Overuse” OR treat* OR therap* OR overtreat* OR undertreat* OR over-treat* OR under-treat* OR screen* OR diagnos* OR detect* OR test OR tested OR testing OR antibiot* OR anti-bacter* OR antibacter* OR antimicrobial OR anti-microbial OR ceftriaxone OR azithromycin OR doxycycline)

#3:

TS=(“Emergency Service, hospital” OR (emergenc* NEAR/2 (depart* OR room* OR service* OR unit* OR ward or wards)))

#4

(#1 AND #2 AND #3)

#### Cochrane*

**Date searched:** February 26, 2021

**Results:** 160

*There was no date filter for this search. The date range was inadvertently left off.

#1:

[mh “Chlamydiaceae Infections”] OR [mh “Chlamydia trachomatis”] OR [mh Chlamydia] OR [mh Chlamydiaceae] OR [mh Gonorrhea] OR [mh “Neisseria gonorrhoeae”] OR [mh “Sexually Transmitted Diseases”] OR [mh “Sexually Transmitted Diseases, Bacterial”] OR (gonococ* OR gonorrh* OR trachomatis OR chlam?di*):ti,ab,kw OR (“sexually transmitted” NEAR/2 (disease* OR infection*)):ti,ab,kw OR (STD OR STDs OR STI OR STIs):ti,ab,kw

#2:

[mh “Medical Overuse”] OR [mh diagnosis] OR [mh “mass screening”] OR (treat* OR therap* OR overtreat* OR undertreat* OR over-treat* OR under-treat* OR screen* OR diagnos* OR detect* OR test OR tested OR testing):ti,ab,kw OR [mh “Anti-Bacterial Agents”] OR [mh ceftriaxone] OR [mh ^azithromycin] OR [mh doxycycline] OR (antibiot* OR anti-bacter* OR antibacter* OR antimicrobial OR anti-microbial OR ceftriaxone OR azithromycin OR doxycycline):ti,ab,kw

#3:

[mh “Emergency Service, hospital”] OR emergenc*:ti OR (emergenc* NEAR/2 (depart* OR room* OR service* OR unit* OR ward OR wards)):ti,ab,kw

#4

(#1 AND #2 AND #3)

#### PsycInfo

**Date searched:**

**Results:** 45

**Dates: 2010-date of search**

**#1:**

(DE “Gonorrhea”) OR (DE “Sexually Transmitted Diseases”) OR ((TI gonococ* OR AB gonococ*) OR (TI gonorrh* OR AB gonorrh*) OR (TI trachomatis OR AB trachomatis) OR (TI chlam#di* OR AB chlam#di*)) OR ((TI “sexually transmitted” OR AB “sexually transmitted”) N2 ((TI disease* OR AB disease*) OR (TI infection* OR AB infection*))) OR ((TI STD OR AB STD) OR (TI STDs OR AB STDs) OR (TI STI OR AB STI) OR (TI STIs OR AB STIs))

#2:

DE “drug therapy” OR (DE “diagnosis”) OR ((TI treat* OR AB treat*) OR (TI therap* OR AB therap*) OR (TI overtreat* OR AB overtreat*) OR (TI undertreat* OR AB undertreat*) OR (TI over-treat* OR AB over-treat*) OR (TI under-treat* OR AB under-treat*) OR (TI overuse OR AB overuse) OR (TI screen* OR AB screen*) OR (TI diagnos* OR AB diagnos*) OR (TI detect* OR AB detect*) OR (TI test OR AB test) OR (TI tested OR AB tested) OR (TI testing OR AB testing)) OR (DE “Antibiotics”) OR ((TI antibiot* OR AB antibiot*) OR (TI anti-bacter* OR AB anti-bacter*) OR (TI antibacter* OR AB antibacter*) OR (TI antimicrobial OR AB antimicrobial) OR (TI anti-microbial OR AB anti-microbial) OR (TI ceftriaxone OR AB ceftriaxone) OR (TI azithromycin OR AB azithromycin) OR (TI doxycycline OR AB doxycycline))

#3:

(DE “Emergency Services”) OR TI emergenc* OR ((TI emergenc* OR AB emergenc*) N2 ((TI depart* OR AB depart*) OR (TI room* OR AB room*) OR (TI service* OR AB service*) OR (TI unit* OR AB unit*) OR (TI ward OR AB ward) OR (TI wards OR AB wards)))

#4

(#1 AND #2 AND #3)

